# Multicenter, Randomized, Open-Label, Comparative Study of Therapeutic Efficacy, Safety and Tolerability of BNO 1030 application in the technology of delayed prescription of antibiotics in patients with severe acute tonsillitis

**DOI:** 10.1101/2021.03.10.21253255

**Authors:** V. I. Popovych, I. V. Koshel, O. N. Malofiichuk, L. I. Pyletska, O. A. Semeniuk, O. V. Martynnyk, R. N. Orlovska, O. I. Leta

**Affiliations:** Ivano-Frankivsk National Medical University; Institute of Postgraduate Education, IFNMU; Regional Clinical Hospital. OF Gerbachevsky ZhOR; Ivano-Frankivsk Regional Clinical Hospital; Danylo Halytsky Lviv National Medical University; KNP 1st city polyclinic of Lviv; The first private child of the polyclinic. M. Ivano-Frankivsk

**Keywords:** severe acute tonsillitis, delayed antibiotic therapy, BNO 1030

## Abstract

Acute bacterial tonsillitis occurs in 20 –30 % of immunocompetent children; however, the frequency of antibacterial drug prescriptions reaches up to 90 %. Delayed antibiotic prescription is recommended by current guidelines. The study objective was to determine the efficacy of phytoneering extract BNO 1030 in the technology of delayed antibiotic prescription in patients with severe acute tonsillitis.

**Methods:** In the multicenter, randomized, open-label, comparative study, 182 out of 200 randomized children with acute tonsillitis aged 6 –12 years completed the study. Evaluation criteria: a reduced severity of sore throat when swallowing and at rest, throat irritation at rest, hyperemia of the tonsils assessed by a physician according to a 4-point scale at each visit compared to Visit 1, dynamics of self-assessment of general well-being, intensity of sore throat and difficulty swallowing according to a 10-point visual analogue scale, frequency of antibiotic prescriptions, therapeutic benefit from BNO 1030 in days.

**Results:** The use of phytotherapeutic medicinal product BNO 1030 in addition to the standard symptomatic treatment of severe acute tonsillitis provides a clinically significant, adequate reduction in the symptom severity assessed by a physician at V2 (p < 0.005). There are significant differences in the patient’s self-assessment of the symptoms from treatment Day 2 (p < 0.005). This allows to significantly reduce the duration of systemic antipyretic administration (p < 0.005). In the first days of treatment, when a decision on delay of antibiotic prescription is made, a therapeutic benefit in two days in patients of the treatment group was observed compared to the control group. The use of BNO 1030 in patients with severe acute tonsillitis significantly reduces, by 43.7 % or 2.3 times, the need for prescribing antibiotic therapy as part of the technology of delayed antibiotic prescription (p < 0.005). During treatment, no side effects and complications of the disease were recorded.

**Conclusion:** BNO 1030 is a safe and effective medicinal product for the treatment of severe acute tonsillitis in children aged 6 –12 years. It provides a significant therapeutic benefit when administered in addition to standard symptomatic therapy and reduces the irrational antibiotic prescription.

**Trial registration:** ClinicalTrials.gov Identifier: NCT04537819 https://clinicaltrials.gov/ct2/show/NCT04537819?term=ATi-2&draw=2&rank=1

## Introduction

Acute tonsillitis (AT) (J03.0 –J03.9) is a very common infectious disease characterized by inflammation of the tonsils. It occurs in all age groups but more often at the age of 5–15 years and accounts for about 5 % of all visits to the physician [1]. Most cases are viral. The most common cause is adenovirus and Epstein-Barr virus. Acute bacterial tonsillitis occurs in immunocompetent children in 20 –30 % of cases, in adults — in 5–15 %, and its cause is group A beta-hemolytic streptococcus (GABHS) [2]. It should be noted that there are no reliable symptoms of bacterial tonsillitis. To determine the indications for antibiotic therapy, patients are stratified according to the Centor-McIsaac scale. However, even with a high total score (> 3), the probability of detecting GABHS in a smear ranges from 30 % to 50 % [3, 4]. Thus, antibiotic therapy is not indicated in most AT cases. At the same time, the frequency of prescribing antibacterial drugs in acute tonsillitis ranges from 50 % to 90 % [5, 6]. Irrational antibiotic prescription in viral tonsillitis, in particular associated with the Epstein-Barr virus, is related to a high incidence of severe, generalized rashes involving limbs [7]. Irrational antibiotic prescription is one of the main causes of the global problem of antibiotic resistance, since inadequate antibiotic therapy is prescribed on average in 50 % of cases worldwide according to WHO data [4, 8, 9].

Delayed antibiotic therapy is one of the strategies to reduce irrational antibiotic prescription in uncomplicated acute respiratory infections [10, 11]. Delayed antibiotic prescriptions show almost the same patient satisfaction rate as antibiotic prescription (86 % versus 91 %). The technology of delayed prescription has reduced the frequency of antibiotic therapy to 31 % and does not lead to an increase in the number of streptococcus-associated complications [5, 6].

Expectations of antibiotic prescription are highest in patients with sore throat and fever. Nevertheless, their prescription has practically no effect on these symptoms, if they are not associated with a bacterial infection [12]. According to current recommendations, paracetamol, often in combination with ibuprofen, is successfully used for symptomatic treatment in patients with sore throat and fever [13, 14]. Several studies have shown that fever inhibits viral replication and has a stimulating effect on the patient’s immune system. Therefore, antipyretics may even prolong the duration of infectious diseases [15 –19]. In addition, paracetamol has been associated with the risk of some potentially dangerous side effects and unintentional overdose in children [20 –22].

In light of the targeted effect on the reduction in the clinical manifestations of tonsillitis, the use of herbal medicinal products could be interesting, since according to studies, phytotherapy for inflammatory diseases of the pharynx is prescribed by 28 % of doctors [23]. The most studied preparations in this respect are *Echinacea* and *Pelargonium sidoides* preparations. However, randomized studies have not shown their efficacy in patients with acute tonsillitis [2, 24]. In clinical practice, a combination phytoneering aqueous extract of BNO 1030 consisting of seven medicinal herbs is used, namely: Marshmallow root (Radix Althaeae), Cammomile flowers (Flores Chamomillae), Horstail herb (Herba Equiseti), Walnut leaves (Folia Jungladis), Yarrow herb (Herba Millefolii), Oak bark (Cortex Quercus), Dandelion herb (Herba Taraxaci), known as Imupret® (known in some countries as Tonsilgon® N). The components of the drug provide antiviral, antibacterial, anti-inflammatory and immunomodulatory effects [25, 26, 27]; indications for use are “treatment of upper respiratory tract diseases (tonsillitis, pharyngitis, laryngitis) and the prevention of complications in viral respiratory infections”. Clinical studies in children have shown that the additional use of the phytoneering drug BNO 1030 (Imupret®) for the treatment of acute tonsillitis significantly reduces the clinical symptoms of tonsillitis, improves the assessment of the patients’ general well-being and quality of life, reduces the use of NSAIDs and the overall duration of treatment with no side effects [28, 29]. However, in the scientific literature, there are no reports on efficacy studies of Imupret® with delayed antibiotic prescription in patients with acute rhinosinusitis which comply with GCP standards.

The objective of this study was to evaluate the efficacy of the additional use of phytoneering extract BNO 1030 in the technology of delayed antibiotic prescription in patients with severe acute tonsillitis versus standard symptomatic therapy according to clinical guidelines [2].

## 2. Materials and methods

### 2.1. Trial design

Multicenter, Randomized, Open-Label, Comparative Study of Therapeutic Efficacy, Safety and Tolerability of Imupret application in the technology of delayed prescription of antibiotics in patients with severe acute tonsillitis was conducted in six outpatient institutions in Ukraine from July 2019 to January 2020. The study was conducted in accordance with the GCP standards and the Declaration of Helsinki. In addition, the study was approved by the Ethics Committee at all study sites. The parents of each child gave their written consent to participate in the study.

### 2.2. Participants

210 outpatients were enrolled in this study. 200 outpatients aged 6–12 years diagnosed with acute bacterial tonsillitis were randomized and divided into two groups: the treatment group of patients who received BNO 1030 in addition to standard therapy and the control group of patients who received standard symptomatic therapy. The treatment group (n = 100) included 47 (47 %) boys and 53 (53 %) girls (average age 8.61 ± 1.994), the control group included 43 (43 %) boys and 57 (57 %) girls (average age 8.51 ± 1.789).

Diagnostic and differential diagnostic criteria for acute tonsillitis were carried out in accordance with the DEGAM recommendations provided in clinical guidelines adopted in Europe and Ukraine [2]. A clinical diagnosis of AT was made based on the presence of such symptoms as sore throat at rest and in swallowing, hyperaemia and swelling with possible plaque on the tonsils, cervical lymphadenitis and fever. The diagnosis of severe acute tonsillitis was made in the case of 4-5 points according to the McIsaac Scale.

### Inclusion criteria

— Male and female outpatient subjects aged 6-12 years diagnosed with acute tonsillitis
— The possibility to start treatment within 72 hours after the disease symptoms occur; 4–5 points according to the McIsaac Scale
— Willingness and ability of the patient and (or) parents to comply with the requirements of the Study Protocol
Signed informed consent

### Withdrawal criteria

— The decision of the patient and/or parents to discontinue participation in the study and withdrawal of written informed consent
— Loss of contact with the patient, individual intolerance to the investigational medicinal product and/or to the reference treatment regimen, the occurrence of serious and/or unforeseen adverse events/reactions in a patient during the study
— Significantly reduced general condition, the development of complications of the underlying disease, which in the physician’s opinion require patient’s withdrawal from the study
— Patient’s violation of the procedures provided by the Protocol

### Exclusion criteria

— 1 to 3 points according to the McIsaac Scale; indications for immediate initiation of systemic antibiotic therapy
— Suspected infectious mononucleosis (clinically)
— The use of systemic antibacterial or antifungal drugs, systemic glucocorticosteroids, cytostatics in the last 14 days
— Intolerance or increased individual sensitivity to any of the components of the investigational medicinal product and the reference treatment regimen

The patients of two groups were of similar sex, age, clinical manifestations of the disease (p < 0.05).

### 2.3. Interventions

From the time of randomization, patients of two groups have been keeping to a sparing diet; irritating factors (physical and chemical) were eliminated; benzydamine spray 3–4 times a day, paracetamol (if case of pain, hyperthermia). Patients of the treatment group additionally received BNO 1030, drops per os, from one batch in the following dosage strength: children aged 6 –11 years — 15 drops 6 times a day; children above 11 years — 25 drops 6 times a day.

BNO 1030, drops per os, is a standardized alcoholic aqueous extract. *Active substances:* 100 g drops contain 29 g of an alcoholic aqueous extract (extracting agent: ethanol 59 % (V/V) made from the following medicinal plants:

|  |  |
| --- | --- |
| Marshmallow root (Radix Althaeae) | 0.4 g; |
| Cammomile flowers (Flores Chamomillae) | 0.3 g; |
| Horstail herb (Herba Equiseti) | 0.5 g; |
| Walnut leaves (Folia Jugladis) | 0.4 g; |
| Yarrow herb (Herba Millefolii) | 0.4 g; |
| Oak bark (Cortex Quercus) | 0.2 g; |
| Dandelion herb (Herba Taraxaci) | 0.4 g; |

*Excipients*: Ethanol 19 % (V/V), purified water.

Name and address of the manufacturer: Bionorica SE, Kerschensteinerstrasse, 11 – 15, 92318, Neumarkt, Germany.

The drug is registered in Ukraine and available over-the-counter (OTC). Therefore, formulation, manufacturing process, packaging and labelling of the dru g comply with GMP and current national requirements of Ukraine. A detailed description of all aspects of the quality and safety of BNO 1030 drops is part of the corresponding product characteristics. Approved indications for use are the treatment of upper respiratory tract diseases (tonsillitis, pharyngitis, laryngitis) and the prevention of complications and recurrences in viral respiratory infections.

ENT practitioners with experience of at least 5 years were involved in the study.

### 2.4. Outcome measures

All data were evaluated by a physician at the beginning of the study and at three follow-up visits within 10 days (Table 1).

**Table 1.**
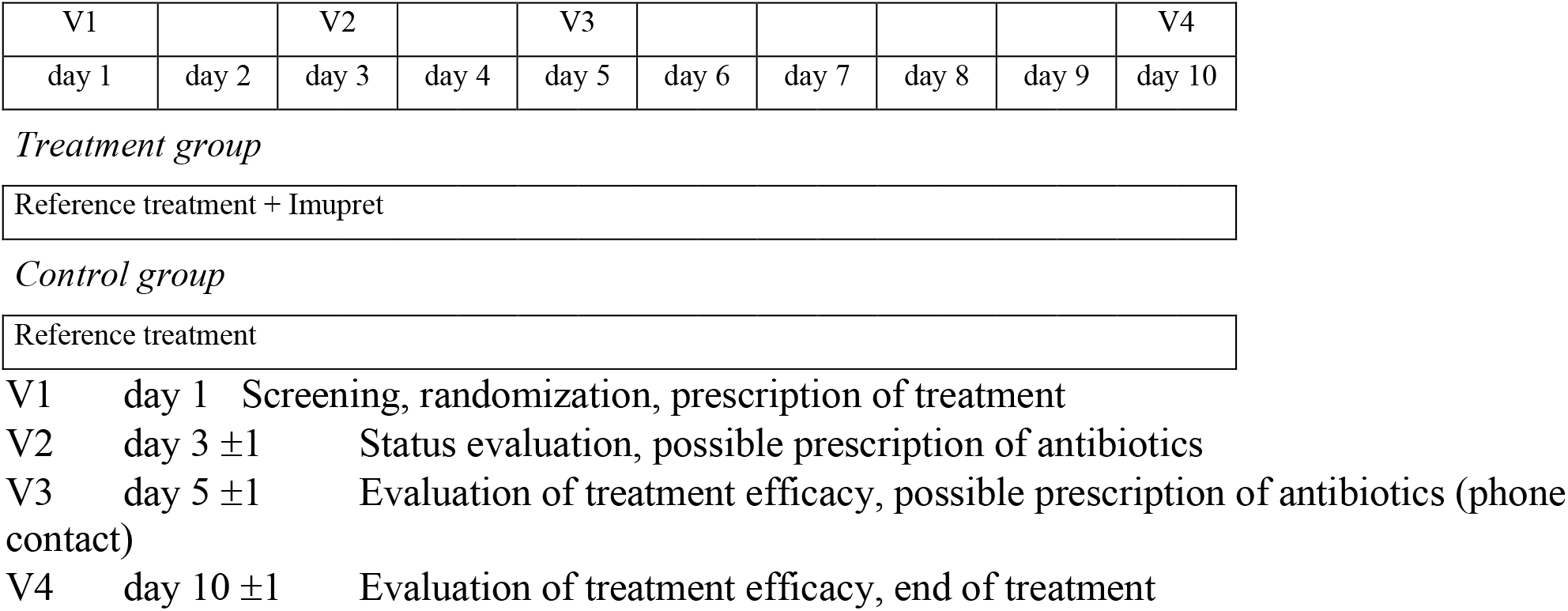
Schedule of assessments.

Schedule of assessments
| V1 |  | V2 |  | V3 |  |  |  |  | V4 |
| --- | --- | --- | --- | --- | --- | --- | --- | --- | --- |
| day 1 | day 2 | day 3 | day 4 | day 5 | day 6 | day 7 | day 8 | day 9 | day 10 |

*Treatment group*
|  |
| --- |
| Reference treatment + Imupret |

*Control group*
|  |
| --- |
| Reference treatment |

|  |  |  |
| --- | --- | --- |
| V1 | day 1 | Screening, randomization, prescription of treatment |
| V2 | day 3 ±1 | Status evaluation, possible prescription of antibiotics |
| V3 | day 5 ±1 | Evaluation of treatment efficacy, possible prescription of antibiotics (phone contact) |
| V4 | day 10 ±1 | Evaluation of treatment efficacy, end of treatment |

#### Schedule of assessments

The physician evaluated the symptoms included in the scale of local manifestations of tonsillitis: sore throat when swallowing and at rest, throat irritation at rest, hyperaemia of the tonsils. All symptoms were evaluated according to a 4-point scale (0 — no symptoms, 1 — mild, 2 — moderate, 3 — severe/pronounced). In the diary, the patient daily assessed the general well-being, intensity of sore throat and difficulty swallowing according a ten-point visual analogue scale. At Visits 2 and 3, the physician assessed the patient’s condition and made a decision on the need to prescribe the antibiotic therapy.

The key efficacy endpoint was: a reduced severity of each symptom (complaint) included into the scales of tonsillitis manifestations, up to 1 point or less, the absence of indications for antibiotic therapy prescription.

Secondary efficacy endpoints: dynamics in a reduced severity of symptoms of the disease assessed according a point scale at Visits 2, 3 and 4 compared with Visit 1; a decrease in the total score (the sum of points for each symptom) according to the local scale manifestations of tonsillitis at Visits 2, 3 and 4 compared with Visit 1; a decrease in temperature in the armpit at Visits 2 and 3 compared with Visit 1; the patient’s self-assessment of the quality of life (daily); duration of NSAID administration.

### 2.5. Sample size

The clinical study has been developed to obtain a reliable description of clinical efficacy of active (additional) use of BNO 1030 compared to the standard treatment only. Depending on findings, several trial descriptive and statistical evaluations were performed so that a biometric estimate of the sample size is not required. However, in order to guarantee a sufficient sample size for data analysis, the sample size N = 200 was chosen. Patients were sorted in a 1:1 ratio.

### 2.6. Randomization

The clinical part of the randomized study is open, without a blinding procedure. Subjects are randomized to one of two possible treatments according to the basic randomization list. Randomization was performed using the software [StatSoft is a random number generator]. Randomization was performed for each patient who signed an informed consent.

### 2.7. Statistical methods

In order to analyse homogeneity of groups, descriptive statistics methods were used for description of the baseline condition of the treatment and control group (for quantitative parameters — n, mean arithmetic, median, standard deviation, minimum and maximum values; for qualitative parameters — incidence and share as %). Verification of normality of data distribution in groups was performed for quantitative parameters using Shapiro-Wilk test. If the data in groups showed normal distribution according to certain parameters, the groups were compared by these parameters using Student’s test for independent samples. Otherwise (if the data distribution was different from normal), comparison of groups was performed using Mann-Whitney test. For categorical parameters, the groups were compared using Pearson’s chi-squared test or Fisher’s exact test.

For analysis of efficacy, descriptive statistics parameters were calculated in each group (n, mean arithmetic, median, standard deviation, minimum and maximum values) for all visits in accordance with patients’ examination scheme. Analysis of dynamics of the said parameters in each group was performed using two-way analysis of variance (ANOVA) according to the following scheme: “Visit” factor is fixed (levels: Visit 1… Visit n); “Subjects” factor is random. Results of the subsequent visits were compared against the data of Visit 1 via contrast analysis using simple contrasts. Comparison between groups in dynamics of tested parameters was performed by differences dTi = (TVisit n−TVisit 1) of assessed parameters using Mann-Whitney test. The level of confidence for Shapiro-Wilk test was accepted equal to 0.01, and for the rest of the criteria it was accepted equal to 0.05.

The analysis was performed in software environment IBM SPSS 22.0

## 3. Results

### 3.1. Study sample

210 patients aged 6-12 years were enrolled in the study (Fig. 1).

**Fig. 1.**
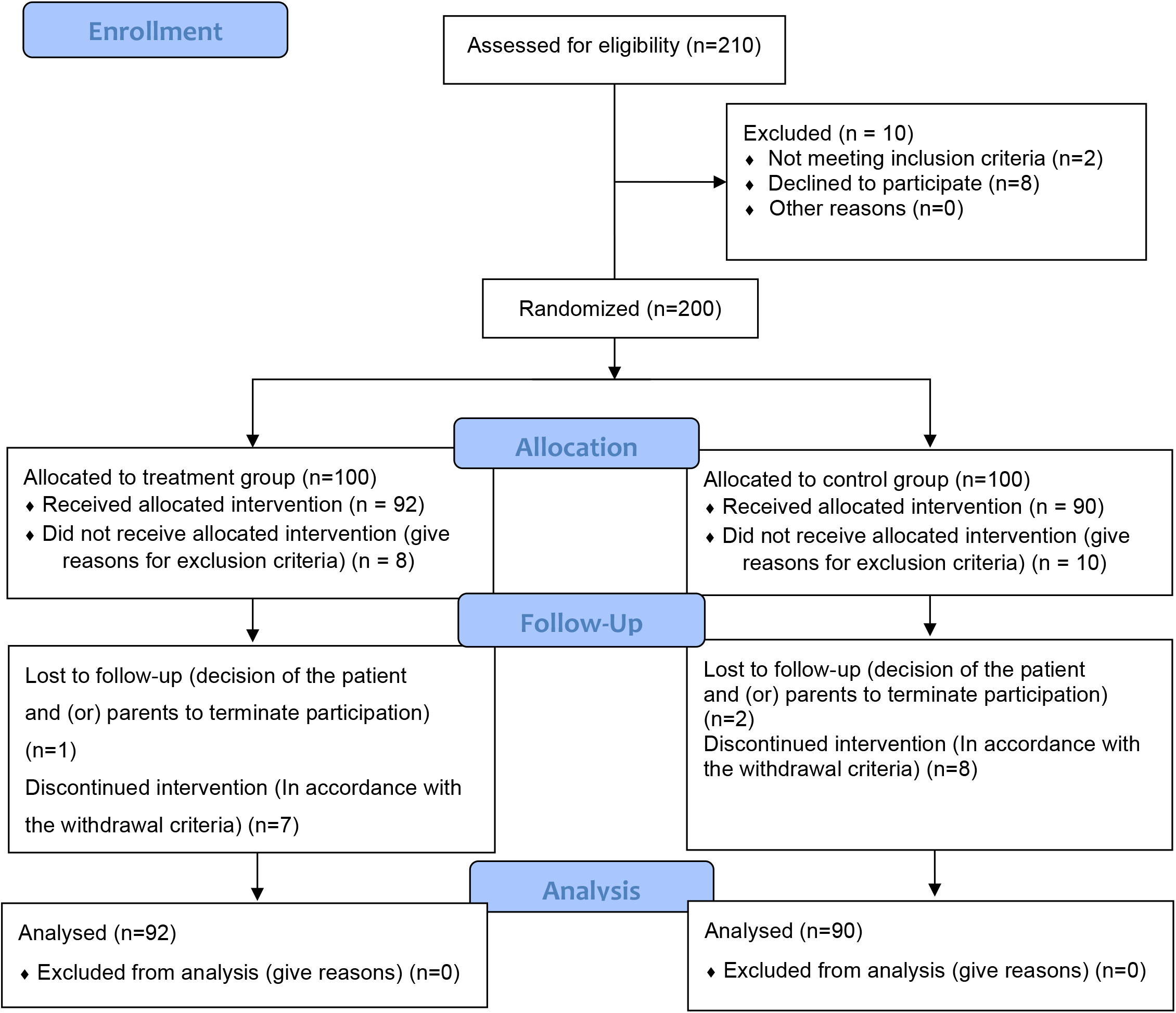
Patients included in screening, randomization and excluded from the study Of the 210 patients enrolled, 10 (4.7 %) were not included in the study. The reason was non-compliance with the study inclusion criteria: age non –compliance (*n* = 2) and the unwillingness of a patient and/or his/her parents to comply with the protocol requirements (*n* = 8). The remaining 200 patients were randomized to either the control group (n = 100) or the treatment group (n = 100). 18 patients (9.0 %) were excluded from the study. The reason was the presence of exclusion criteria (decision to discontinue participation in the study) (n = 10) in the control group and (n = 8) in the treatment group. Thus, from July 2019 to January 2020, 182 (91 %) out of the 200 randomized patients (n = 92 in the treatment group) and (n = 90 in the control group) completed the study and were analysed.

The gender distribution in the groups was as follows: out of 100 patients in the treatment group, 47 (47 %) were boys and 53 (53 %) were girls; out of 100 patients in the control group, 43 (43 %) were boys and 57 (57%) were girls. There were slightly more girls than boys (55 % vs. 45 %) among patients enrolled in the study. According to the age criterion, the groups were homogeneous: the average age of patients was 8.61 **±** 1.994 in the treatment group and 8.51 ± 1.789 in the control group.

In general, there were no significant differences in demographic characteristics among patients from the treatment and control groups at baseline (Day 1) (P < 0.05).

Table 2 shows the comparative characteristics of the treatment and control groups according to the total score of the tonsillitis severity in points using the McIsaac Scale.

**Table 2.**

| Parameter | Group | Statistical indicators |  |  |  |
| --- | --- | --- | --- | --- | --- |
| | | n | M±SD | p-value | Homogeneity of the groups* |
| McIsaac scale | Treatment | 100 | 4.45 ± 0.500 | P = 0.569 | Homogeneous |
| | Control | 100 | 4.41 ± 0.494 | | |

\* The conclusion is drawn at the significance level of 0.05

All patients had severe tonsillitis since the symptom severity in all patients included in the study was > 4 points: 4.41 ± 0.494 in the control group and 4.45 ± 0.500 in the treatment group. There were no differences in the severity of tonsillitis between patients at baseline (Day 1) (p < 0.05).

### 3.2. Outcomes and estimation

The main clinical manifestation most typical for a patient with acute tonsillitis is a sore throat. Table 3 shows the results of the physician assessment of symptom severity of sore throat when swallowing and at rest, as well as throat irritation in patients of both groups on three follow-up visits.

**Table 3.** Dynamics of the symptom severity of acute tonsillitis depending on the group.

| Parameter | Statistical indicators |  |  |  |  |  |  |  |  |  |
| --- | --- | --- | --- | --- | --- | --- | --- | --- | --- | --- |
|  | Visit | Treatment group |  |  | Control group |  |  | Comparison of the groups |  |  |
|  |  | n | M±SD | Regression (%) | n | M±SD | Regression (%) | dT | p-value | Significant difference * |
| Pain when swallowing | V1 | 100 | 2.69 ± 0.526 | - | 100 | 2.59 ± 0.605 | - | - | - | - |
|  | V2 | 99 | 1.65 ± 0.733 | -38.7 | 98 | 2.06 ± 0.831 | -20.5 | dT<sub>2</sub> | 0.000 | <u>Sign</u> |
|  | V4 | 92 | 0.16 ± 0.532 | -94.1 | 90 | 0.71 ± 0.874 | -72.6 | dT<sub>4</sub> | 0.000 | <u>Sign</u> |
| Sore throat alone | V1 | 100 | 2.43 ± 0.671 | - | 100 | 2.47 ± 0.611 | - | - | - | - |
|  | V2 | 99 | 1.30 ± 0.788 | -46.5 | 98 | 1.84 ± 0.765 | -25.5 | dT<sub>2</sub> | 0.000 | <u>Sign</u> |
|  | V4 | 92 | 0.11 ± 0.425 | -95.5 | 90 | 0.48 ± 0.619 | -80.6 | dT<sub>4</sub> | 0.000 | <u>Sign</u> |
| Throat irritation alone | V1 | 100 | 2.16 ± 0.813 | - | 100 | 1.96 ± 0.844 | - | - | - | - |
|  | V2 | 99 | 1.13 ± 0.680 | -47.7 | 98 | 1.52 ± 0.899 | -22.4 | dT<sub>2</sub> | 0.000 | <u>Sign</u> |
|  | V4 | 92 | 0.12 ± 0.409 | -94.4 | 90 | 0.47 ± 0.601 | -76.0 | dT<sub>4</sub> | 0.000 | <u>Sign</u> |
| Redness of the tonsils | V1 | 100 | 2.39 ± 0.723 | - | 100 | 2.42 ± 0.727 | - | - | - | - |
|  | V2 | 99 | 1.41 ± 0.729 | -41.0 | 98 | 1.94 ± 0.895 | -19.8 | dT<sub>2</sub> | 0.000 | <u>Sign</u> |
|  | V4 | 92 | 0.33 ± 0.697 | -86.2 | 90 | 0.96 ± 0.947 | -60.3 | dT<sub>4</sub> | 0.000 | <u>Sign</u> |
| * The conclusion is drawn at the significance level of 0.05 |  |  |  |  |  |  |  |  |  |  |

At baseline (V1), there were no significant differences in the sore throat pain intensity at rest, pain in swallowing and throat irritation between patients in the treatment and control groups (p < 0.05). Since initially the groups did not differ in assessment of the symptom severity, the individual differences dT2 = (V2-V1) and dT4 = (V4-V1) were calculated for each patient by each parameter. In the course of treatment at V2, the intensity of these symptoms was significantly less pronounced in the treatment group compared to the control group (significant differences, p > 0.05). Between V2 and V3, there was a further decrease in symptom severity in both groups. Significant differences between the groups were observed even at the end of treatment (p > 0.05).

Table 4 shows the results of self-assessment of the condition within 10 days of treatment for three symptoms: general well-being, sore throat and difficulty swallowing (from 0 to 10 points for each symptom).

**Table 4.** Dynamics of tonsillitis symptoms according to patients’ self-assessment.

| Parameter | Statistical indicators |  |  |  |  |  |  |  |  |  |
| --- | --- | --- | --- | --- | --- | --- | --- | --- | --- | --- |
|  | Day | Treatment group |  |  | Control group |  |  | Comparison of the groups |  |  |
|  |  | n | M±SD | Regression (%) | n | M±SD | Regression (%) | dT | p-value | Significant difference * |
| Self-assessment of well-being | D1 | 100 | 7.23 ± 1.524 | - | 100 | 7.10 ± 1.475 | - | - | - | - |
|  | D2 | 99 | 6.25 ± 1.710 | -13.6 | 98 | 6.76 ± 1.632 | -4.8 | dT<sub>2</sub> | 0.000 | Sign |
|  | D3 | 99 | 5.07 ± 1.797 | -29.9 | 97 | 6.12 ± 1.883 | -13.8 | dT<sub>3</sub> | 0.000 | Sign |
|  | D4 | 98 | 3.96 ± 2.056 | -45.2 | 96 | 5.27 ± 2.085 | -25.8 | dT<sub>3</sub> | 0.000 | Sign |
|  | D5 | 95 | 3.20 ± 2.061 | -55.7 | 95 | 4.49 ± 2.310 | -36.8 | dT<sub>5</sub> | 0.000 | Sign |
|  | D6 | 90 | 2.26 ± 1.696 | -68.7 | 89 | 4.06 ± 1.979 | -42.8 | dT<sub>6</sub> | 0.000 | Sign |
|  | D7 | 87 | 1.52 ± 1.593 | -79.0 | 85 | 3.49 ± 1.919 | -50.8 | dT<sub>7</sub> | 0.000 | Sign |
|  | D8 | 83 | 1.02 ± 1.370 | -85.9 | 80 | 2.99 ± 1.831 | -57.9 | dT<sub>8</sub> | 0.000 | Sign |
|  | D9 | 83 | 0.51 ± 1.086 | -92.9 | 77 | 2.29 ± 1.669 | -67.7 | dT<sub>9</sub> | 0.000 | Sign |
|  | D10 | 83 | 0.25 ± 1.948 | -96.5 | 77 | 1.69 ± 1.489 | -76.2 | dT<sub>10</sub> | 0.000 | Sign |
| Self-esteem sore throat | D1 | 100 | 6.71 ± 2.006 |  | 100 | 6.77 ± 1.711 |  |  | - | - |
|  | D2 | 99 | 5.82 ± 2.002 | -13.3 | 98 | 6.41 ± 1.858 | -5.3 | dT<sub>2</sub> | 0.000 | Sign |
|  | D3 | 99 | 4.56 ± 2.016 | -32.0 | 97 | 5.86 ± 1.995 | -13.4 | dT<sub>3</sub> | 0.000 | Sign |
|  | D4 | 98 | 3.64 ± 2.022 | -45.8 | 96 | 5.11±2.051 | -24.5 | dT<sub>3</sub> | 0.000 | Sign |
|  | D5 | 95 | 2.86 ± 1.916 | -57.4 | 95 | 4.43 ± 2.191 | -34.6 | dT<sub>5</sub> | 0.000 | Sign |
|  | D6 | 90 | 2.04 ± 1.779 | -69.6 | 89 | 3.94 ± 1.979 | -41.8 | dT<sub>6</sub> | 0.000 | Sign |
|  | D7 | 87 | 1.45 ± 1.468 | -78.4 | 85 | 3.37 ± 1.853 | -50.2 | dT<sub>7</sub> | 0.000 | Sign |
|  | D8 | 83 | 0.98 ± 1.370 | -85.4 | 80 | 2.81 ± 1.801 | -58.5 | dT<sub>8</sub> | 0.000 | Sign |
|  | D9 | 83 | 0.47 ± 0.980 | -93.0 | 77 | 2.08 ± 1.692 | -69.3 | dT<sub>9</sub> | 0.000 | Sign |
|  | D10 | 83 | 0.18 ± 0.665 | -97.3 | 77 | 1.48 ± 1.498 | -78.1 | dT<sub>10</sub> | 0.000 | Sign |
| Self-assessment difficulty swallowing | D1 | 100 | 6.39 ± 2.006 |  | 100 | 6.39 ± 2.049 |  |  | - | - |
|  | D2 | 99 | 5.55 ± 2.214 | -13.1 | 98 | 6.02 ± 2.082 | -5.8 | dT<sub>2</sub> | 0.000 | Sign |
|  | D3 | 99 | 4.36 ± 2.169 | -31.8 | 97 | 5.55 ± 2.250 | -13.1 | dT<sub>3</sub> | 0.000 | Sign |
|  | D4 | 98 | 3.34 ± 2.192 | -47.7 | 96 | 4.72 ± 2.209 | -26.1 | dT<sub>3</sub> | 0.000 | Sign |
|  | D5 | 95 | 2.61 ± 2.054 | -59.2 | 95 | 4.09 ± 2.343 | -36.0 | dT<sub>5</sub> | 0.000 | Sign |
|  | D6 | 90 | 1.82 ± 1.669 | -71.5 | 89 | 3.55 ± 2.094 | -44.4 | dT<sub>6</sub> | 0.000 | Sign |
|  | D7 | 87 | 1.21 ± 1.542 | -81.1 | 85 | 3.07 ± 2.046 | -52.0 | dT<sub>7</sub> | 0.000 | Sign |
|  | D8 | 83 | 0.72 ± 1.300 | -88.7 | 80 | 2.54 ± 1.935 | -60.3 | dT<sub>8</sub> | 0.000 | Sign |
|  | D9 | 83 | 0.31 ± 0.896 | -95.1 | 77 | 1.94 ± 1.681 | -69.6 | dT<sub>9</sub> | 0.000 | Sign |
|  | D10 | 83 | 0.19 ± 0.818 | -97.0 | 77 | 1.43 ± 1.517 | -77.6 | dT<sub>10</sub> | 0.000 | Sign |

\* The conclusion is drawn at the significance level of 0.05

At the beginning of the study (Day 1), the results of the assessment performed by the patients were similar in both groups. Since initially the groups did not differ in the self-assessed severity of symptoms, individual differences dT1 = (Day 2-Day 1) were calculated. …. dT10 = (Day 10-Day 1) by each parameter. Starting from Day 2, a more pronounced regression of main complaints was observed in the treatment group (more than 13 %) compared to the control group (less than 6 %) (p > 0.05). A significantly more pronounced regression of main complaints was observed in the treatment group on all consecutive days of follow-up period till the end of the follow-up period (p > 0.05 in all cases). In general, the results of patients’ self-assessment of symptoms in dynamics correspond to the results of the physicians’ assessment (statistically significant difference between the groups).

Improvement of local symptoms of acute tonsillitis and general well-being leads to a decrease in the amount of systemic non-steroidal anti-inflammatory drugs taken. The dynamics of NSAID administration was assessed (Table 5). The last drug administration date was taken into account.

**Table 5.** The dynamics of systemic NSAID administration depending on the group.

| Day/Group | 1 | 2 | 3 | 4 | 5 | 6 | 7 | 8 | 9 | 10 |
| --- | --- | --- | --- | --- | --- | --- | --- | --- | --- | --- |
| Treatment group<br>(n=100) | 70 | 55 | 25 | 6 | 2 | 1 | 0 | 0 | 0 | 0 |
|  | 70.7 % | 55.6 % | 25.5 % | 6.3 % | 2.2 % | 1.15 % | 0.0 % | 0.0 % | 0.0 % | 0.0 % |
| Control group<br>(n=100) | 70 | 62 | 50 | 29 | 13 | 10 | 3 | 0 | 0 | 0 |
|  | 70.7 % | 63.9 % | 52.1 % | 30.5 % | 14.6 % | 11.8 % | 3.8 % | 0.0 % | 0.0 % | 0.0 % |
| p-value | 0.946 | 0.294 | 0.0003 | 0.0001 | 0.003 | 0.005 | 0.116 | 1.000 | 1.000 | 1.000 |
| Conclusion* | - | - | Sign | Sign | Sign | Sign | - | - | - | - |
| *(-) There are no significant differences between the groups according to NSAIDs<br>(Sign) There are significant differences between the groups according to NSAIDs |  |  |  |  |  |  |  |  |  |  |

There was a significant difference in NSAID administration between patients of the treatment and control groups starting from Day 3 of treatment (p > 0.05). The duration of antipyretic administration by patients of the treatment group was 2.36 ± 1.649 days, which is less than in the control group: 3.12 ± 0.494. The difference is significant (p = 0.014).

The groups were compared according to the number of patients who received antibacterial drugs (Fig. 2).

**Figure 2.**
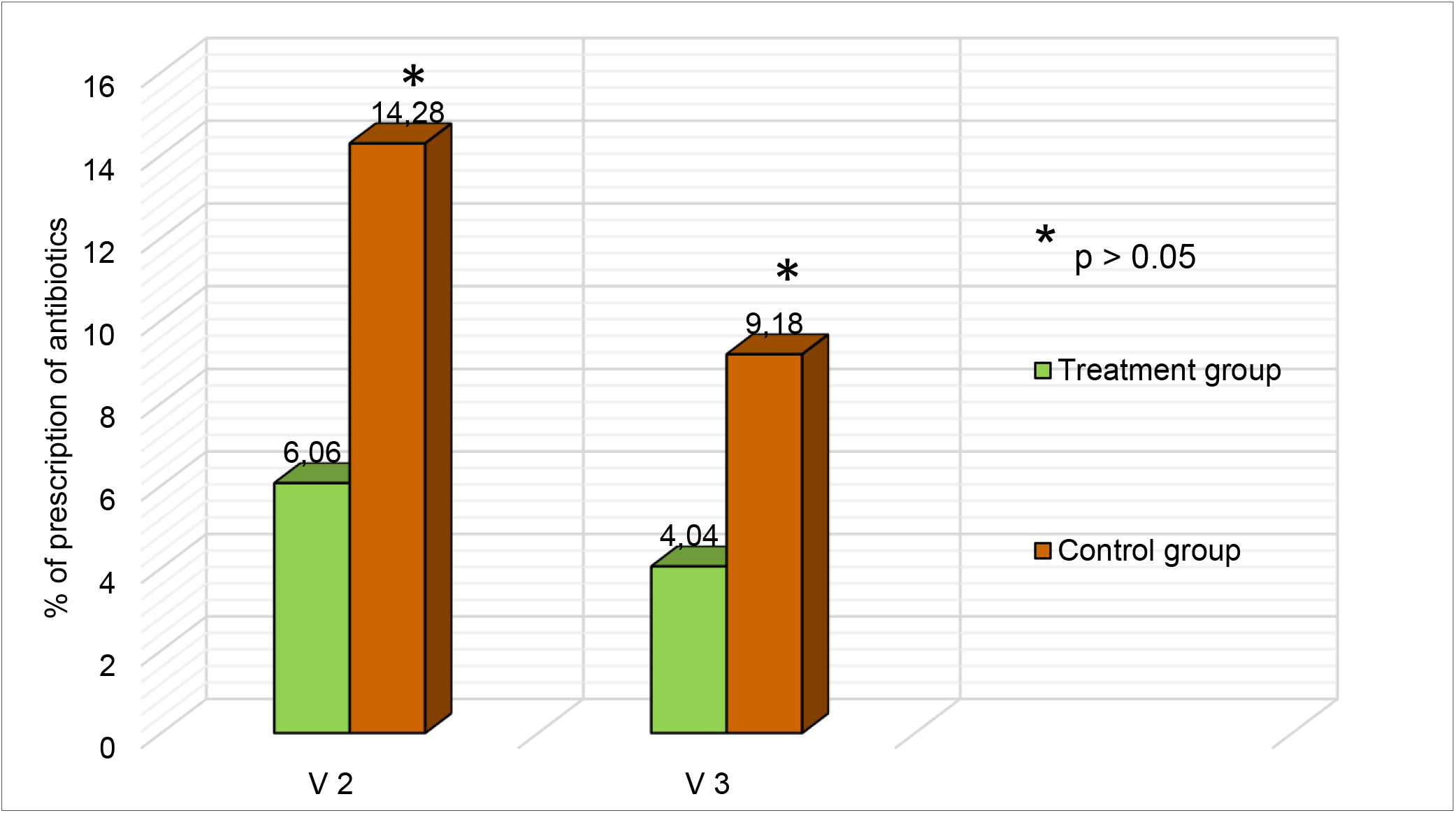
Antibiotic prescription in the groups.

As can be seen from Figure 2, antibiotics were prescribed at V2 to 6 (6.06 %) patients in the treatment group and 14 (14.28 %) patients in the control group. Two days later (V3), antibiotics were additionally prescribed to 4 (4.04 %) patients in the treatment group and 9 (9.18 %) patients in the control group. Thus, in accordance with the technology of delayed prescription, antibiotic treatment was prescribed to 10 (10.1 %) patients in the treatment group and 23 (23,46) patients in the control group. The difference between the groups was significant (P = 0.0223).

### 3.3. Safety and tolerability

An analysis of the tolerability assessment findings showed that the treatment was well tolerated or very well tolerated in all cases. In the course of treatment, no significant side effects, as well as streptococcus-associated complications requiring additional treatment or drug withdrawal, were registered in any patient.

## 4. Discussion

Acute bacterial tonsillitis occurs in immunocompetent children in 20 –30 % of cases [2]. Current guidelines recommend symptomatic agents (topical and systemic NSAIDs) for symptom relief, primarily sore throat. For antibiotic prescription, patients are stratified using the Centor-McIsaac Scale. When the condition is assessed in > 3 points, immediate antibiotic prescription is recommended only if streptococcus is verified. In the absence of verification, antibiotics are prescribed using the technology of delayed prescription [10, 11]. However, the frequency of antibacterial drug prescriptions in severe acute tonsillitis reaches 90 % [5, 6]. In terms of using a delayed antibiotic strategy, initial treatment should be highly effective. Many researchers are of the opinion that with low confidence in the regression of tonsillitis symptoms during treatment, there is always a question about the antibiotic prescription both among physicians and the desire for antibiotic therapy among patients themselves or their parents [12]. According to the design, our study enrolled patients with diagnostic criteria for severe acute tonsillitis. The symptom severity according to the McIsaac Scale in the treatment group was 4.45, in the control group — 4.41 points. It has been demonstrated that the use of the standardized phytoextract BNO 1030 in addition to standard symptomatic therapy for severe acute tonsillitis has proven therapeutic benefits. Patients in the treatment group demonstrated a clinically significant reduction in the symptom severity assessed by the physician as early as V2 (p > 0.005). There were significant differences in the patient’s self-assessment of general well-being, the severity of sore throat and difficulty swallowing according a 10-point scale starting as early as Day 2 (D2) of treatment (p > 0.005). An important and interesting conclusion of the study is that due to a more pronounced regression of sore throat, as well as an improvement in general well-being, patients in the BNO 1030 group discontinued systemic antipyretics earlier. The duration of their administration was 2.36 ± 1.649 days compared to 3.12 ± 0.494 in the control group. The difference in the antipyretic administration was significant starting from Day 3 of treatment (p > 0.05).

Thus, the patients of the treatment group showed a significant “therapeutic benefit” in comparison with the control group starting from Day 2 of treatment. As part of the technology of delayed prescription, this is very important since the decision to prescribe antibiotics is made after assessing the dynamics of symptom regression on Day 3–5 of treatment.

As known, it is difficult for a physician to adhere to the technology of complete refusal to prescribe antibiotics in acute tonsillitis, especially when assessing the condition according to the Centor-McIsaac Scale in three or more points. That is why the delayed prescription is considered in patients with severe tonsillitis. In our study, the technology of delayed antibiotic prescription was used in 23 (23 .46 %) patients in the control group. The number of prescriptions corresponds to the average incidence of bacterial tonsillitis [2]. Antibiotic therapy was prescribed to 10 (10.12 %) patients of the treatment group (p > 0.05). Thus, an important conclusion of the study is that the use of BNO 1030 in patients with severe acute tonsillitis significantly reduces, by 43.7 % or 2.3-fold, the need for antibacterial therapy when using the technology of delayed antibiotic prescription.

At the same time, according to the literature, the delayed prescription tactics are not widespread and the frequency of antibacterial drug prescriptions in acute tonsillitis reaches up to 90 % [5, 6]. According to the DESCARTE study, the difference in the level of immediate and delayed prescriptions in patients with a Centor score of > 3 points is very modest, not more than 13 %, and not statistically significant [30]. The proven efficacy of BNO 1030 is an important argument to reduce the urge of physicians and patients to immediate antibiotic prescription.

The proven high efficacy of acute tonsillitis treatment in the treatment group in terms of severe regression of symptoms in the first two-three days will make it possible to more widely implement the delayed antibiotic prescription strategy and greatly reduce the number of irrational antibiotic prescriptions.

The efficacy of BNO 1030, shown in this study, is generally confirmed by the earlier study results in patients with acute non-bacterial tonsillitis [28]. When BNO 1030 was administered, a “therapeutic benefit” in four days was obtained on Day 10 of treatment compared to the control group. The strength of the present study is the symptom-based stratification of patients of 4 –5 points according to the McIsaac Scale, which corresponds to the concept of “severe acute tonsillitis”. It is the diagnosis of severe tonsillitis that is the main reason for irrational antibiotic prescription. The groups of randomized patients, homogeneous in clinical manifestations, made it possible to draw reasonable conclusions on comparative treatment results. The “therapeutic benefit” at the time of the decision on the delayed antibiotic prescription (V2, V3) is more than two days in patients of the treatment group. This reflects a significant advantage in assessing the condition and rationale for prescribing antibiotics as part of the technology of delayed prescription.

The limitation of this study is the lack of placebo control. However, the comparison was carried out during treatment according to clinical recommendations, which provide for the mandatory prescription of symptomatic therapy only with the use of NSAIDs [2, 13, 14]. Consequently, all the differences in treatment results can be attributed to the clinical effects of BNO 1030.

## 5. Conclusions

It was shown that the additional use of the phytoneering drug BNO 1030 for the treatment of severe acute tonsillitis significantly reduces the clinical symptoms, improves the assessment of patients’ general well-being and quality of life starting from Day 2 and reduces the use of NSAIDs from Day 3 of treatment. This makes it possible to reduce antibiotic prescriptions by 43.7 % or 2.3-fold as part of the technology of delayed prescription. The inclusion of the drug in the treatment regimen may be recommended for patients with severe acute tonsillitis.

The direction for future research is to study the efficacy of the drug as an adjunct to antibacterial therapy in patients with acute bacterial tonsillitis.

## Data Availability

All the data can be provided by the corresponding author.

https://clinicaltrials.gov/ct2/show/NCT04537819?term=ATi-2&draw=2&rank=1

